# Monogenic causes of Premature Ovarian Insufficiency are rare and mostly recessive

**DOI:** 10.1101/2022.11.21.22282589

**Authors:** Saleh Shekari, Stasa Stankovic, Eugene J. Gardner, Gareth Hawkes, Katherine A. Kentistou, Robin N. Beaumont, Alexander Mörseburg, Andrew R. Wood, Gita Mishra, Felix Day, Julia Baptista, Caroline F. Wright, Michael N. Weedon, Eva Hoffmann, Katherine S. Ruth, Ken Ong, John R. B. Perry, Anna Murray

## Abstract

Premature ovarian insufficiency (POI) affects 1% of women and is a leading cause of infertility. It is often considered to be a monogenic disorder, with pathogenic variants in ∼100 genes described in the literature. We sought to systematically evaluate the penetrance of variants in these genes using exome sequence data in 104,733 women from the UK Biobank, 2,231 (1.14%) of whom reported natural menopause under the age of 40. In the largest study of POI to date, we found limited evidence to support any previously reported autosomal dominant effect. For nearly all heterozygous effects on previously reported POI genes we were able to rule out even modest penetrance, with 99.9% (13,699/13,708) of all identified protein truncating variants found in reproductively healthy women. We found evidence of novel haploinsufficiency effects in several genes, including *TWNK* (1.54 years earlier menopause, *P*=1.59*10^−6^) and *SOHLH2* (3.48 years earlier menopause, *P*=1.03*10^−4^). Collectively our results suggest that for the vast majority of women, POI is not caused by autosomal dominant variants either in genes previously reported or currently evaluated in clinical diagnostic panels. We suggest that the majority of POI cases are likely oligogenic or polygenic in nature, which has major implications for future clinical genetic studies, and genetic counselling for families affected by POI.

## Introduction

Premature ovarian insufficiency (POI) is the loss of ovarian activity and permanent cessation of menstruation occurring before the age of 40^1^. It represents a major cause of female infertility, affecting 1 in 100 women^1-4^. Some POI cases are syndromic, in which POI accompanies other phenotypic features, such as in Turner’s syndrome. Genetic causes of POI have been reported in 1-10% of cases while other causes include autoimmune and iatrogenic^5-7^. Approximately 50-90% of POI cases are idiopathic^8,9^, 10-30% of those being familial, suggesting a genetic basis. Furthermore, heritability estimates of menopausal age from mother-daughter pairs range from 44% to 65%^10,11^ and there is a six times increased risk of early menopause in daughters of affected mothers^12,13^. A genetic diagnosis can provide important information to families about the risks of POI as well as the aetiology of the condition.

More than 100 monogenic causes of POI have been reported, where a single genetic variant is sufficient to cause the phenotype, with approximately half showing an autosomal dominant (AD) inheritance pattern (e.g. *BNC1, FANCA* and *NOBOX*). Variants in other genes are described as being inherited in an autosomal recessive (AR) manner, requiring both copies of the gene to be disrupted in order to cause the phenotype (eg. *HFM1, LARS2* and *MCM8*). In addition to the autosomal genes, X chromosome genes have long been suggested to play an essential role in the maintenance of ovarian development and function, with X chromosome structural variants representing about 13% of POI cases in some published series^8,14,15^.

More recently, GWAS have identified ∼300 common genetic variants associated with population variation in timing of menopause^11,16^. These studies have provided evidence that some POI cases may be polygenic in nature^11^, where women inherit large numbers of common alleles associated with earlier menopause that, when combined with other risk factors, could push them into the extreme end of the phenotypic distribution.

With decreasing cost and improved analytical pipelines, whole exome sequencing (WES) is increasingly being used in clinical settings as a powerful diagnostic tool, including for POI^17-21^. However, the reported evidence for causal POI genes and variants is inconsistent, often based on small numbers of families or individuals, and with variable degree of functional validation^21^. As genetic testing becomes more widespread in both clinical and non-clinical settings, there is an increasing need to better understand the phenotypic consequences of finding variants in these genes, to help ensure appropriate advice and treatment is offered to women. Therefore, we aimed to assess the penetrance of variants in genes previously reported to cause POI, in a general population study. We focused on the POI genes that are part of the Genomics England diagnostic gene panel for POI, an expert reviewed and publicly available panel database, which we additionally supplemented with literature-reported POI genes. Our results indicate that the reported autosomal dominant (AD) causes of POI are likely to be either only partially penetrant or not pathogenic. Furthermore, we conclude that most cases of menopause under 40 years are likely to be multifactorial.

## Results

### Heterozygous damaging variants do not often cause POI

The Genomics England POI Panel App (version 1.67) includes 67 validated genes rated as either ‘GREEN’ (high level of evidence for disease association), ‘AMBER’ (moderate evidence) or ‘RED’ (not enough evidence). We also identified a further 38 genes reported as being causal for POI. We classified these 105 genes according to the reported mode of inheritance (**Supplementary Table 1**). We then identified genetic variants in these 105 putative POI genes using WES data available in 104,733 UK Biobank post-menopausal female participants of European genetic ancestry^22^, of which 2,231 reported age at natural menopause (ANM) below the age of 40. HC-PTVs were found in 100 genes, but never only in the cases: there were 41 women with menopause under 40 years (ANM range: 27-39, mean ANM: 36.4, SD=3.2) who had a high confidence protein truncating variant (HC-PTV) in at least one of the 40 genes reported to be autosomal dominant, but these variants were also detected in 1,817 women with ANM over 40 years (ANM range: 40-63, mean ANM: 50.4, SD=3.9). For three of the 40 POI genes (*BMPR1A, FOXL2* and *NR5A1)* there were no HC-PTVs carriers in either cases or controls, but for all 37 genes with HC-PTVs, the median ANM for those with heterozygous loss of function (LOF) alleles was between 45 and 56 years (**Figure 1, Supplementary Table 3**).

**Figure 1:**
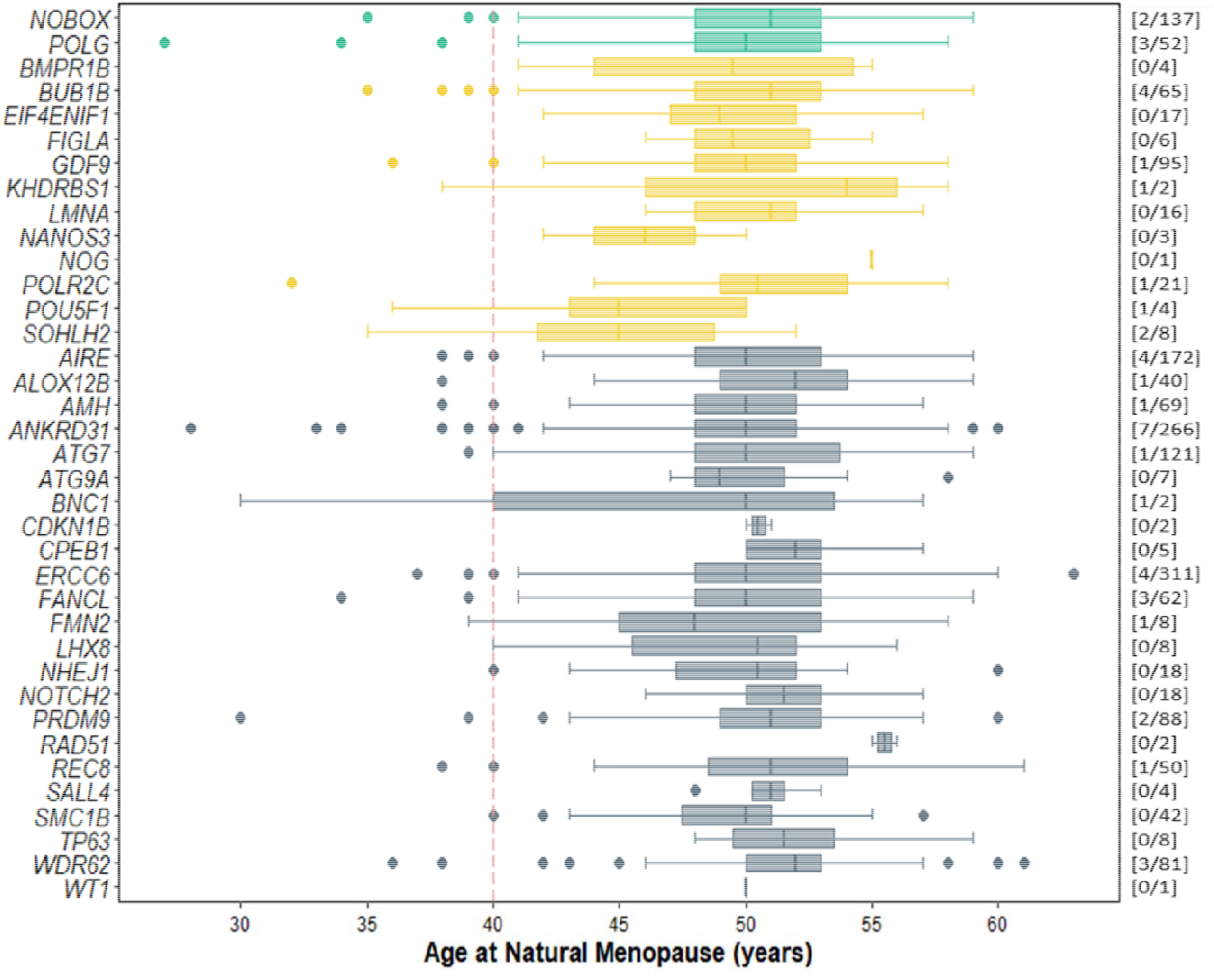
Age at natural menopause in women with HC-PTVs in POI genes reported to have an autosomal dominant pattern of inheritance. *Genes are coloured by the strength of evidence for POI in either the Genomics England Panel App (Green or Amber; no HC-PTV were detected in red genes) or our own manual curation of the POI literature (grey; N = 23). The number of women with ANM <40 (cases) compared to >40 years (controls) are shown in brackets on the right Y axis [cases/controls].* In the plot, the boxes show the values of the lower quartile, median and upper quartile; the whiskers show the most extreme value within a distance of 1.5 times of the interquartile range from the lower and upper quartiles, respectively; outliers are shown as individual points.

The intolerance for individual genes to harbour protein truncating variation, also known as genic ‘constraint’, has previously been linked to reproductive success^23^. Our results demonstrate that the majority of AD POI genes (26/40, 67.5%) have limited evidence of being under strong selective constraint (pLI ≤ 0.9) as assessed by gnomAD^24^, which further supports that these genes are unlikely to play an important role for reproductive success.

Next we tested individual variants in the 40 AD genes that have been previously reported to be pathogenic for POI (**Supplementary Table 4**). There were 153 variants reported, of which 126 were predicted to be missense and, of these, 58 (46%) were detected in our study with 37 only found in controls. A further 20 missense variants were found both in women with ANM under 40 years and controls, and only one missense variant was found only in cases (NM_002693.3:c.2828G>A {p.Arg943His} in *POLG*]); however, the burden tests of all HC-PTVs or deleterious missense variants in *POLG* were not associated with menopause timing (*P*=0.7 and *P*=0.05, respectively; **Supplementary Table 5**). Therefore, while the variant in cases alone could have a gain of function or dominant negative effect, the finding is also consistent with chance. Having tested reported ‘pathogenic’ missense variants in the 40 AD genes, we tested all missense variants with MAF<0.1% in UK Biobank. We next collated a broader set of 17,374 rare missense variants in the 40 AD genes, including 2,740 with CADD score >25 (**Supplementary Figure 1, Supplementary Table 8**) and 1,120 with REVEL score >0.7 (**Supplementary Figure 2, Supplementary Table 9**). We identified no robust associations with ANM for any of these individual variants (all were *P*>3.11*10^−4^ and so above our threshold for multiple testing of all missense variants with AC>5; 0.05/4,737=1.06*10^−5^). These results support our previous observation that POI genes are generally not pathogenic in the heterozygous state.

Due to the relatively small number of protein truncating variants (PTVs) found within individual genes, to try to increase our statistical power to find any association with POI we considered the aggregated effect of all PTVs with similar proposed genetic architecture across all putative POI genes. This included a test for: (1) AD only genes (N=38), (2) autosomal recessive (AR) only genes (N=57), (3) genes with both AD and AR inheritance (N=2), and (4) all 105 POI genes. None of the tests were associated with ANM at P<0.05, in either a generalised linear model or STAAR Omnibus statistical models^25^ (Methods; **Supplementary Table 11**).

### No evidence of haploinsufficiency as a cause of POI

Of the 105 reported monogenic POI genes assessed in our study, 57 were reported to show AR inheritance and a further eight were X-linked. We were unable to evaluate recessive effects as we identified only two women with homozygous HC-PTVs: one with a PTV in *SOHLH1* (NM_001101677.2:c.346-1G>A) with menopause at 45 years and one in *AIRE* (NM_000383.4:c.967_979del {p.Leu323SerfsTer51}) who reported menopause in her 20s. Furthermore, we were unable to identify compound heterozygotes. Instead, by considering HC-PTV allele frequencies in our analyses, we would expect 0.003% of individuals (∼4 in the current study) to be homozygous or compound heterozygous for a high-confidence LOF variant in any of the 105 POI genes. This is likely a conservative estimate given we might expect POI genes to be less tolerant than other genes to deleterious alleles as these would impact reproductive fitness. Based on frequencies of gene knockout carriers in gnoMAD^26^, we estimate that even if all genes in the genome were true recessive causes of POI (and thus not detected by our study), the population prevalence of carrying a gene knockout would be 100 times smaller than the observed prevalence of POI.

We next hypothesised that there may be an effect on ANM in heterozygous carriers of deleterious variants in these POI recessive genes. In total we identified 122 carriers of HC-PTVs in the 65 recessive or X-linked genes among cases with ANM < 40 years, but also 5,585 carriers among controls (**Supplementary Table 3**). However, there was no evidence that haploinsufficiency of any recessive POI gene is sufficient to cause POI (**Supplementary Figure 3**).

Finally, we assessed whether protein-coding variation in any of the 105 monogenic POI genes altered ANM within the normal range. In gene burden tests we grouped genetic variants with MAF < 0.1% into three functional categories: (1) HC-PTVs, (2) missense variants with CADD score ≥ 25, and (3) a combination of 1 and 2, termed ‘damaging’ variants. For 100 of the 105 POI genes, we did not find an association with ANM (P<1.6*10^−4^; P=0.05/(3 tests × 105 genes) (**Supplementary Table 5**). For two AR genes, we have previously reported an effect on ANM: *BRCA2* (*P*=2.6*10^−8^; beta:1.32 years earlier ANM [95% CI: −1.79, −0.85]) and *HROB* (*P*=4.7*10^−7^; beta: 2.69 years earlier ANM [95% CI: −3.73, −1.65])^27^. There were novel associations with earlier ANM for a further two AD and one AR genes, with at least one of the variant categories passing our threshold for multiple testing (*P*<1.6*10^−4^, **Figure 2**). These were for damaging variants in *TWNK*, a mitochondrial helicase involved in mtDNA replication and repair (*P*=1.59*10^−6^; beta: 1.54 years earlier ANM [95% CI: −2.17,-0.91]; N = 180)^28,29^, *NR5A1*, a key gene for gonadal function (*P*=5.8*10^−8^; beta: 2.04 years earlier ANM [95% CI: −2.79, −1.30]; N = 131)^30^, and *SOHLH2*, a transcription factor involved in both male and female germ cell development and differentiation (*P*=1.03*10^−4^; beta: 3.48 years earlier ANM [95% CI: −5.24, − 1.72]; N = 23)^31,32^.

**Figure 2:**
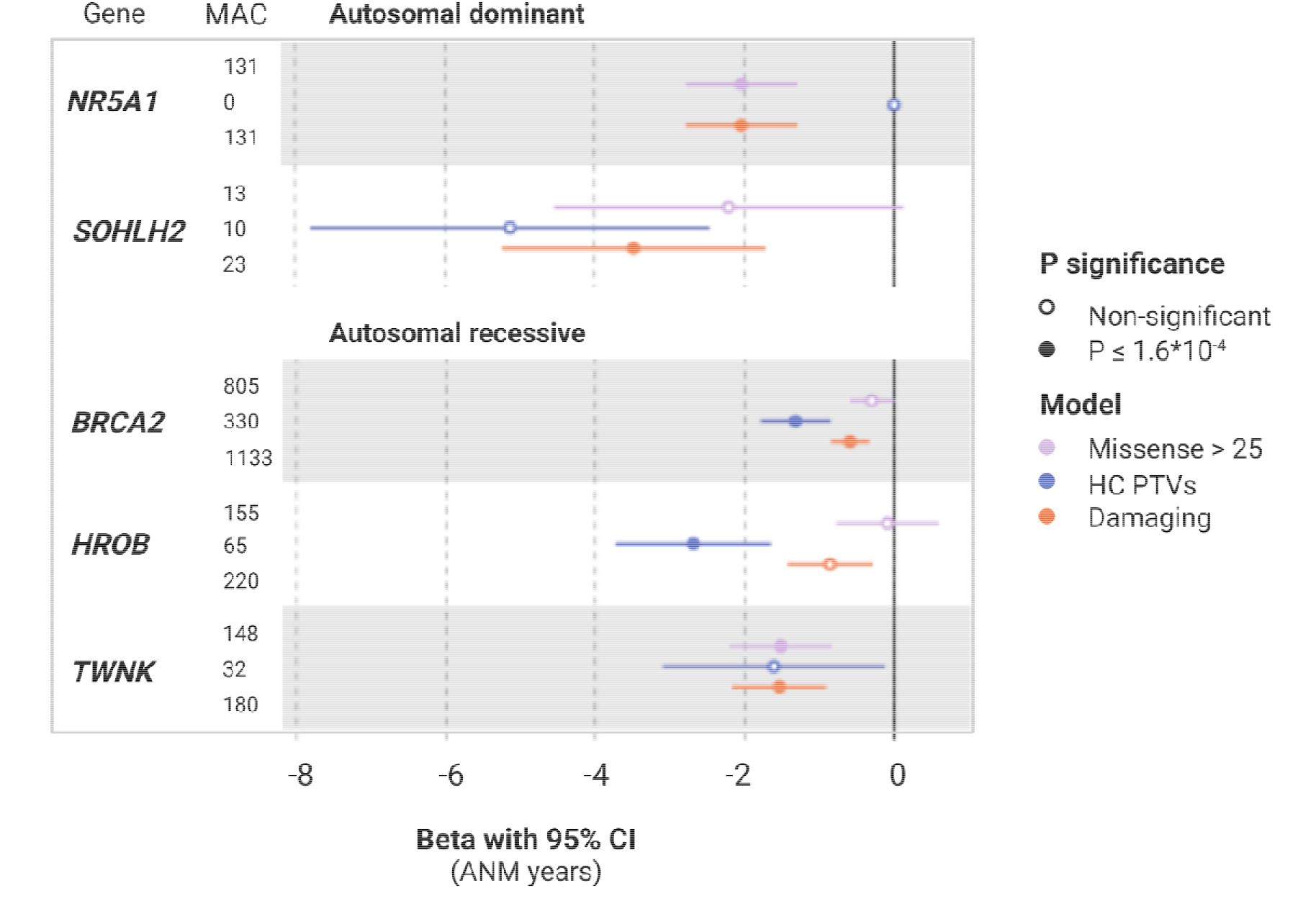
Gene burden associations with age at natural menopause. Results are plotted for genes that passed the Bonferroni corrected threshold for 105 genes, each with 3 masks (P<1.6×10^−4^). There were no HC-PTV carriers for NR5A1.

## Discussion

Many genes in the literature have been reported as monogenic causes of POI and are included in diagnostic panels for clinical use^33^. Our literature review identified 105 putative monogenic POI genes, 67 that were included in the Genomics England open access Panel App resource^34^ and 38 additional reported genes. Of these 105 genes, 40 are reported to be inherited in an AD fashion. Using UK Biobank exome sequence data in 104,733 post-menopausal women, we found no evidence to support heterozygous HC-PTV of any of these genes as a highly penetrant cause of POI; for each gene the average menopause age for carriers of LOF variants was over 40, with the ANM distribution broadly similar to that of non-carriers. This includes two green Panel App genes - *NOBOX* and *POLG* - where 137/139 and 52/55 of the identified PTV alleles, respectively, were found in controls. Our previous work demonstrated that heterozygous LOF of *ZNF518A* has the largest effect in the protein-coding genome on menopause timing ^27^, yet carriers report menopause only 6 years earlier than non-carriers, with only 12% experiencing POI. Taken together, our observations suggest that fully, or even largely, penetrant autosomal dominant effects are likely to cause very few cases of POI.

Although heterozygous LOF variants were not penetrant causes of POI, our study suggests that carrying rare coding variants in five of the POI genes can substantially lower an individual’s menopause age. Besides previously reported *BRCA2* and *HROB*, three genes, including *NR5A1, SOHLH2* and *TWNK*, have not been described as associated with menopause timing in the general population. The effect ranged from 5.13 years earlier menopause for carriers of rare LOF variants in *SOHLH2* to 1.54 years earlier for damaging variants in *TWNK*. We did not identify any heterozygous PTV alleles in *NR5A1*, which is a highly constrained gene (**Supplementary Table 1**). Therefore, the observation that rare missense variants are associated with a ∼2 year reduction in menopause timing suggests that dominant LOF may well be a penetrant, albeit very rare, cause of POI. As for *NR5A1, TWNK* is also on the green gene on the Panel App list, but has a reported recessive pattern that causes syndromic POI (Perrault syndrome) and presents in association with other neurologic symptoms^35^.

Our findings should be interpreted in the context that the published evidence to support causality of genes and variants for POI is highly variable. Guidelines are available for genomic variant interpretation^36^, but many of these genes were reported before such guidelines, making it difficult for non-specialists to interpret the findings. Many studies were based on candidate gene approaches with small numbers of cases or families^18^ and in the absence of large-scale reference data or ancestry match controls. More recent POI studies have used exome sequencing, but often revert to candidate gene approaches with relaxed statistical thresholds when no exome-wide association is identified^17,20,37,38^. Furthermore, when studying individual genomes it is also inherently challenging distinguishing between pathogenic variants and private non-functional variants. Functional studies can be informative, but the design and rationale of such studies can be circular. For example, a DNA damage response (DDR) gene that harbours a private variant may be selected as a reasonable candidate, but the downstream functional work is limited to DDR measures, rather than reproductive or ovarian phenotypes. Future studies that aim to investigate novel genetic causes of POI should focus on approaches that more specifically mimic human biology and physiology. Patient-specific induced pluripotent stem cells (iPSCs) lines might offer an individually targeted genetic model for identification, manipulation and better understanding of reproductive biological pathways.

Our study has assessed one of the largest samples to date of women with menopause before 40. A major strength is the analyses of exome sequence data in over 100,000 women with normal ANM, which provides invaluable data on normal genetic variation in a control population. Identification of alleles at high frequency in these samples provides confidence that they are unlikely to be a penetrant cause of POI. Our study does however have a number of limitations. Firstly, we have not investigated a clinically defined cohort of POI cases and not all women with menopause under 40 would be diagnosed as POI. Furthermore, the UK Biobank is known to disproportionately include healthier participants, which has been shown to be the case for other conditions^39,40^, although this tends to have a greater impact on men^41^ and it is not obvious that having POI would influence participation in the study. While these issues will likely lead to underestimates of any potential effect sizes, they do not explain why previously reported pathogenic variants are overwhelmingly found in women with menopause over 40 years. Secondly, we were able to assess the penetrance only of heterozygous variants but not homozygous or compound heterozygous carriers. We also have not considered complex structural variants or cytogenetic abnormalities, so we make no statement on the penetrance of those. For five genes we did not identify any heterozygous LOF variants in our data so were unable to assess these, although they are unlikely causes of POI given they were not present in over 2000 cases. Third, we predominantly focussed on predicted LOF alleles as the mechanism implied or demonstrated in most studies. It is however possible that some of the literature reported missense variants may act in a gain of function or dominant negative manner such that they have more severe effects than LOF variants. Whilst potentially true of a small number, this is unlikely to be widespread given no highly penetrant effects were seen in the 58 individual literature reported missense variants that we assessed, or in our burden tests for predicted damaging missense variants by CADD and REVEL. Finally, our study is specific to individuals of European ancestry. While the frequencies of many variants vary between populations, the functional impacts of LOF variants should be widely applicable.

In conclusion, our findings imply that monogenic causes of POI are unlikely for the vast majority of cases. Given our observed results for genes with a dominant mode of inheritance, we advise caution in interpreting reported recessive effects, although we predict this will be by far the most common cause of monogenic POI. Rather than representing a biologically distinct condition, we suggest that POI is part of a continuous distribution of ovarian ageing. Where women are in this distribution is likely determined by a continuum of multiple risk factors, where the sum of many independent genetic and non-genetic risk factors place women into the tail of the phenotypic distribution. This notion is also supported by our recent work reporting that women with the top 1% of a polygenic risk score, comprising common ANM-reducing alleles, have a five-fold increased risk of POI compared to the median^11^. Collectively, our findings suggest that POI should be considered a genetically complex trait for which genetic testing for monogenic causes is unlikely to be fruitful. Future efforts should address this genetic complexity in the development of new diagnostic approaches for POI to minimise potential mis-diagnoses and inappropriate genetic counselling.

## Methods

### Identification of reported POI genes

In order to identify relevant gene candidates reported to cause POI, we initially focused on the POI gene panel available through Genomics England Panel App, publicly accessible virtual panel database (https://panelapp.genomicsengland.co.uk/panels/155/). This panel was selected as the ‘gold standard’ resource as it is the most thoroughly curated one, reviewed by 12 professional clinical geneticists. We considered the following evidence as part of our gene evaluation: (1) Selection and categorisation: inheritance and phenotype, and (2) Number of reviews and gene ranking based on their traffic light system. This includes “RED” genes that do not have enough evidence for the association with the condition and should not be used for clinical interpretation, “AMBER” genes with moderate evidence that should not be <u>yet</u> used for the interpretation, and “GREEN” genes with high level of evidence, which demonstrates confidence that this gene should be used for clinical interpretation (**Supplementary Table 1**). In total, we identified 67 genes: 28 green, 23 amber and 16 red. We reviewed the evidence provided on the Genomics England Panel App webpage for these genes, and identified the specific genetic variants reported as associated with the phenotype (**Supplementary Table 2**).

This list was additionally supplemented with 38 manually curated POI genes reported in the literature. The search was performed using PubMed and Google Scholar, focusing on original articles published up to June 2022. The key word combinations included ‘premature ovarian failure’, ‘primary ovarian insufficiency’, ‘premature ovarian insufficiency’, ‘early menopause’, ‘premature menopause’, ‘POI’, ‘POF’, ‘infertility’, ‘hypergonadotropic hypogonadism’, ‘ovarian dysgenesis’, ‘genetic variants’, ‘sequencing’, and ‘primary amenorrhea’. Studies were also identified by a manual search of original publications described in review articles. Where appropriate, reference lists of identified articles were also searched for further relevant papers. Identified articles were restricted to English language full-text papers. Studies were included according to following criteria: (1) the phenotype of interest was described as POI, primary or secondary amenorrhea, (2) one or more affected individuals for particular causal variant were identified, (3) the focus was on either the autosomes or the X chromosome, (4) genetic variants were discovered by traditional family segregation studies, consanguineous pedigree analysis, unrelated cohort studies on whole exome (WES)/targeted next-generation sequencing data, (5) variant discovery was supported by validation in animal models and/or cell based assays. We excluded studies that: (1) described hypothalamic pituitary adrenal axis and/or puberty related phenotypes, (2) genes that were discovered through genome-wide association studies due to the lack of statistical power as a result of small sample sizes and the challenge to locate causative genes, and finally (3) genes that were discovered through array analysis due to the high inconsistency of the results coming from varied resolution of arrays across studies and thus uncommon replications. We recorded and analysed genes described for either non-syndromic or syndromic POI, however the main focus of our paper was on genes associated with non-syndromic POI. Papers that exclusively reported the role of candidate genes in animal models, were only used as supporting evidence when assessing the functional evaluation of the gene and to guide our conclusions. Following information were extracted from each study: (1) Publication info: PMID, (2) Inheritance: autosomal dominant (AD), autosomal recessive (AR) or X-linked, (3) Sample size: number of the genetic variant carriers, cases versus controls, if reported, and (4) Genetic variant info: genomic position, transcript and protein sequence (**Supplementary Table 2**). If the data were missing from published papers, relevant information was obtained by direct communication with the corresponding authors. In cases where response was not received, the information was recorded as NA. All data were extracted independently by two authors (S. Shekari and S. Stankovic).

Overall, we identified 105 unique POI genes that we classified according to their mode of inheritance. This includes, 67 validated genes rated as either ‘GREEN’ (high level of evidence for disease association), ‘AMBER’ (moderate evidence) or ‘RED’ (not enough evidence) on the Genomics England POI Panel App (version 1.67). We also identified a further 38 genes reported as being causal for POI (**Supplementary Table 1**).

Genes were considered as inherited through the AD pattern if the reported variants in the heterozygous state were sufficient to cause POI, leading to 40 genes in total. Of those, seven were reported to act through the LoF mechanism only, while in 34 genes both LoF and missense genetic alterations caused the phenotype. If variants in both copies of the gene were necessary for the phenotype development the gene was classified as AR (N=57). For two genes (*POLG, REC8*) both dominant and recessive causes were identified and so we investigated them with other AD genes, while seven genes had an X-linked inheritance pattern.

### Constraint metric of pathogenicity

We annotated each gene identified with the Genome Aggregation Database (gnomAD) v2.1.1 predicted constraint metric of pathogenicity to identify genes that are subject to strong selection against PTV variation^24^. The metric encompassed observed and expected variant counts per gene, observed/expected ratio (O/E) and probability of loss of function intolerance (pLI) (**Supplementary Table 1**). In short, observed count represents the number of unique SNPs in each gene (MAF < 0.1%), while expected count relies on a depth-corrected probability prediction model that takes into account sequence context, coverage and methylation to predict expected variant count. The O/E is a continuous measurement that assesses how tolerant a gene is to a certain class of variation. Low O/E value indicates that the gene is under stronger selection for that class of variation. Finally, the pLI score reflects the constraint or intolerance of a given gene to a PTV variation, with a score closer to 1 indicating that the gene cannot tolerate PTV variation.

### UK Biobank Data Processing and Quality Control

To perform rare variant burden analyses described in this study, we accessed Whole Exome Sequencing data (WES) for 454,787 individuals from the UK Biobank study^42^. Details of this study, including data collection and processing, are extensively described elsewhere^43^. Informed consent was provided by all participants. Study approval was received from the National Research Ethics Service Committee North West–Haydock and all study procedures were performed according to the World Medical Association Declaration of Helsinki ethical principles for medical research.

WES data were generated with the IDT xGen Exome Research Panel v1.0, which targeted 39Mbp of the human genome with mean coverage exceeding 20x on 95.6% of sites. The OQFE protocol was used for mapping and variant calling to the GRCh38 reference. Quality control filters applied by UK Biobank were individual and variant missingness <10% and Hardy Weinberg Equilibrium P-value >10^−15^. In addition, we excluded variants with <10X coverage in 90% of the samples that were provided by Backman *et al*. ^42^. We selected variants in the Consensus CDS (CCDS) transcripts and variants were annotated using the Ensembl Variant Effect Predictor^44^ and LOFTEE plugin (https://github.com/konradjk/loftee). Minor allele frequency (MAF) was calculated using PLINK^45^. Furthermore, for homozygous variants, we manually assessed the variants using the Integrative Genomics Viewer^46,47^. Analyses were performed on the UK Biobank Research Analysis Platform (RAP; https://ukbiobank.dnanexus.com/).

### Phenotype derivation

ANM was derived from self-reported questionnaire data as the age at last naturally occurring menstrual period, excluding those with surgical menopause (field 2824 and 3882) or taking hormone replacement therapy (field 3536), as described previously^11^. There were 104,733 female participants with ANM included in our analyses (range 18 to 65 years, mean=50.1, SD=4.5); of whom 2,231 individuals reported ANM under 40 years. During the data collection process, participants who reported ANM under 40 years on the questionnaire were asked to confirm their ANM. For comparisons of variant counts, we identified a control cohort of women with ANM at ≥40 years including those who reported still menstruating (n=192,438). Analyses were performed in Stata:Release 16 on the UK Biobank RAP.

### Primary exome-wide association analysis

In order to perform rare variant burden tests, we used the REGENIE regression algorithm (REGENIEv2.2.4; https://github.com/rgcgithub/regenie). REGENIE implements a generalised mixed-model region-based association test that can account for population stratification and sample relatedness in large-scale analyses. REGENIE runs in 2 steps^48^, which we implemented on the UKBiobank RAP: In the first step, genetic variants are aggregated into gene-specific units for each class of variant called masks: high confidence protein-truncating variants included stop-gain, frameshift, or abolishing a canonical splice site (−2 or +2 bp from exon, excluding the ones in the last exon); non-synonymous (missense) variants with CADD score > 25; damaging that included high confidence protein-truncating variants or/and non-synonymous variants with CADD score >25. The three masks were tested for association with ANM in the second step. As described previously, in our analyses we included individuals identified as European, excluding participants who had subsequently withdrawn from the study and those for whom self-reported sex did not match genetic sex^49^. We applied an inverse normal rank transformation to ANM and included recruitment centre, sequence batch and 40 principal components as covariates. We transformed the effect estimates from our analyses to approximate values in years by multiplying by the standard deviation of ANM in our study cohort (4.53 years). Analyses were performed on the UK Biobank RAP. To identify significant gene associations we Bonferroni corrected P<0.05 for the number of masks (n=3) and genes tested (n=105) giving a significance threshold of *P*<1.6*10^−4^ (*P*=0.05/(3*105)=1.6*10^−4^).

In a similar way, we used REGENIE to test the association of individual genetic variants reported in the literature with ANM. Variants with allele count >5 were tested in an additive model, applying an inverse normal rank transformation to ANM and including recruitment centre, sequence batch and 40 principal components calculated by UK Biobank as covariates. Of 421 uniquely identified variants,182 were present in the UK Biobank.

### Replication analyses

A second analysis team (Cambridge) independently performed analyses of WES data in UK Biobank. The ANM phenotype was derived as described in Stankovic *et al* (2022)^27^. Briefly, a different approach was used to generate the phenotype by handling data from multiple visits and missing data differently to the main method of generating the phenotype. This resulted in 106,973 female individuals for analyses. All manipulations were conducted in R (v4.1.2) on the UK Biobank RAP.

Rare variant burden tests of functional variant categories (defined as for main analyses) were performed using a custom implementation of BOLT-LMM v2.3.6^50^ for the UK Biobank RAP, as described in Stankovic *et al* (2022)^27^. Analyses used a winsorised ANM phenotype, with everyone reporting ANM at younger than 34 years given a value of 34. Analyses were adjusted for age, age^2^, sex, the first ten genetic principal components as calculated in Bycroft *et al*.^*51*^ and study participant exome sequencing batch as a categorical covariate (either 50k, 200k, or 450k).

### Gene-set burden analysis

We ran gene-set burden tests by collapsing the genes of interest and their variants into one unit for analysis. The gene-set burden tests were performed by extending an association testing workflow of applets designed for the UK Biobank RAP for single genes to gene-sets. The RAP association workflow is described in detail in Gardner *et al*, 2022^52^. In total, we conducted four gene-set burden tests, collapsing variants and genes into the following categories: (1 AD only genes (N=38), (2) AR genes (N=57), (3) genes with both AD and AR inheritance (N=2), and (4) all 105 genes (**Supplementary Table 11**).

Briefly, for each of the gene-sets we included variants with MAF < 0.1% that were HC-PTVs as predicted by the LOFTEE tool^24^. For each gene-set we ran two related approaches. Firstly, we implemented a generalised linear model (GLM) using the Python package ‘statsmodels’ ^53^. For the GLM, the number of variant alleles across the gene-set was summed up into a single score under a simple additive model. This score was used as a predictor of the ANM phenotype in a three-step regression.

Secondly, we ran the STAAR method (implemented in R package “STAAR”)^25^. This method corrects for population stratification by including a genetic relatedness matrix (GRM) in the test framework. The GRM used was based on pre-computed autosomal kinship coefficients from Bycroft *et al* ^*51*^. For each STAAR test the genotype information was represented by a single n*p matrix where n was the sample size and p the number of included genetic variants across all genes of interest. For all association tests we corrected for age, age^2^, the first ten genetic principal components provided by Bycroft *et al* ^51^ and study participants WES batch as a categorical covariate.

### Frequency of homozygous or compound heterozygous LOF individuals

We estimated the frequency of homozygous or compound heterozygous HC-PTV individuals for each gene as *F*^2, where *F* is the frequency of individuals with any high-confidence HC-PTV allele with MAF<0.1% in a gene as estimated from the primary analysis (**Supplementary Table 3**). To find the total frequency of individuals with homozygous or compound heterozygous HC-PTVs, we then summed *F*^2 for the 105 POI genes reported in the literature.

The expected frequency of having a gene with a homozygous or compound heterozygous LOF knockout is 6 per billion individuals, based on the median frequency in gnomAD^26^. From this estimate we would expect 1.2 per 10,000 people to carry a homozygous or compound heterozygous LOF knockout in any of the ∼20,000 genes in the genome (20000*6*10^−9^=1.2*10^−4^). Assuming 100% penetrance, the number of genes with a homozygous or compound heterozygous LOF knockout that would be needed to reach the observed 1% frequency of POI in the population (1 per 100 individuals) would be 0.01/6*10^−9^=1.7*10^6^ genes.

## Data Availability

All data produced in the present work are contained in the manuscript

## Acknowledgements

This work was funded by the Medical Research Council (Unit programs: MC_UU_12015/2, MC_UU_00006/2, MC_UU_12015/1, and MC_UU_00006/1). The views expressed are those of the author(s) and not necessarily those of the NIHR or the Department of Health and Social Care. The authors acknowledge the use of the University of Exeter High-Performance Computing facility in carrying out this work, funded by a MRC Clinical Research Infrastructure award (MRC Grant: MR/M008924/1). This study was supported by the National Institute for Health and Care Research Exeter Biomedical Research Centre. For the purpose of open access, the author has applied a Creative Commons Attribution (CC BY) license to any Author Accepted Manuscript version arising. This research was conducted using the UK Biobank Resource under application 9905 (University of Cambridge) and 9072 and 871 (University of Exeter).

Saleh Shekari was supported by the QUEX Institute (University of Exeter, UK and the University of Queensland, Australia). Stasa Stankovic is supported by the Clare Hall Ivan D. Jankovic PhD scholarship from the University of Cambridge. Anna Murray, Caroline Wright and Michael Weedon are supported by the Medical Research Council (MR/T00200X/1). Katherine Ruth is supported by Cancer Research UK [grant number C18281/A29019]. Gareth Hawkes has received funding from the Innovative Medicines Initiative 2 Joint Undertaking under grant agreement No 875534. Gita Mishra is supported by National Health and Medical Research Council Investigator grant (APP2009577). Eva Hoffmann was supported by the ERC (724718-ReCAP), Novo Nordisk Foundation (NNF15COC0016662), the Independent Research Foundation Denmark (0134-00299B), and a grant from the Danish National Research Foundation Centre (6110-00344B).

## Supplementary Figures

**Supplementary Figure 1:**
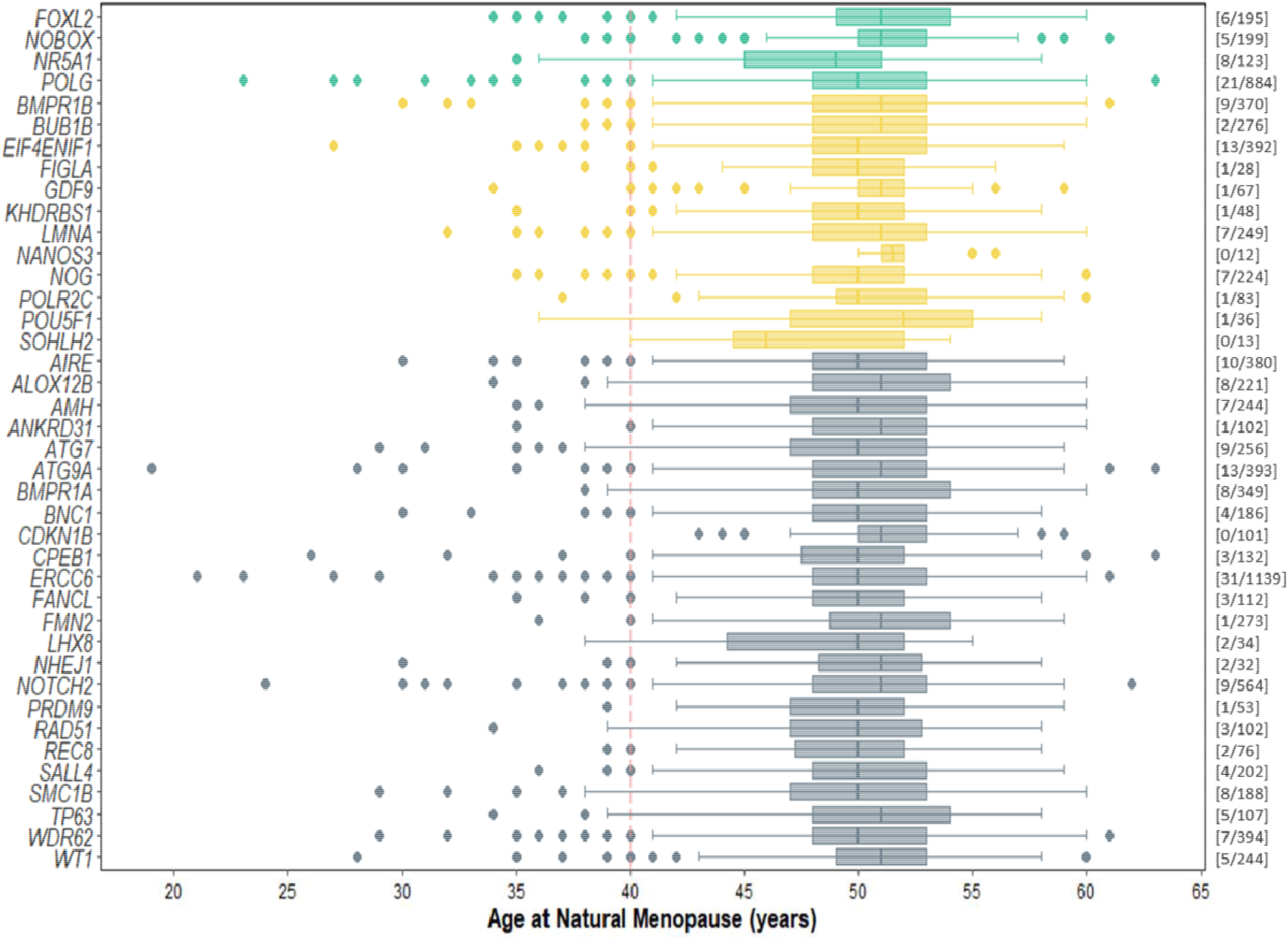
Range of age at natural menopause in carriers of missense variants with CADD score greater than 25 in genes reported to have an autosomal dominant pattern of inheritance. 17 genes were identified as ‘monoallelic’ in Genomics England (GeL) Panel App and are coloured according to the strength of evidence categories: “GREEN”, and “AMBER” **(supplementary table 2)**. In addition,24 genes were reported in the literature to be a likely monogenic cause of POI in the heterozygous state but were not included on the Panel App (coloured grey). The numbers in brackets in the right corner reported as part of each panel represent **[N POI cases/N controls]** of women carrying HC PTVs in each gene. Note: In the plot, the boxes show the values of the lower quartile, median and upper quartile; the whiskers show the most extreme value within a distance of 1.5 times of the interquartile range from the lower and upper quartiles, respectively; outliers are shown as individual points.

**Supplementary Figure 2:**
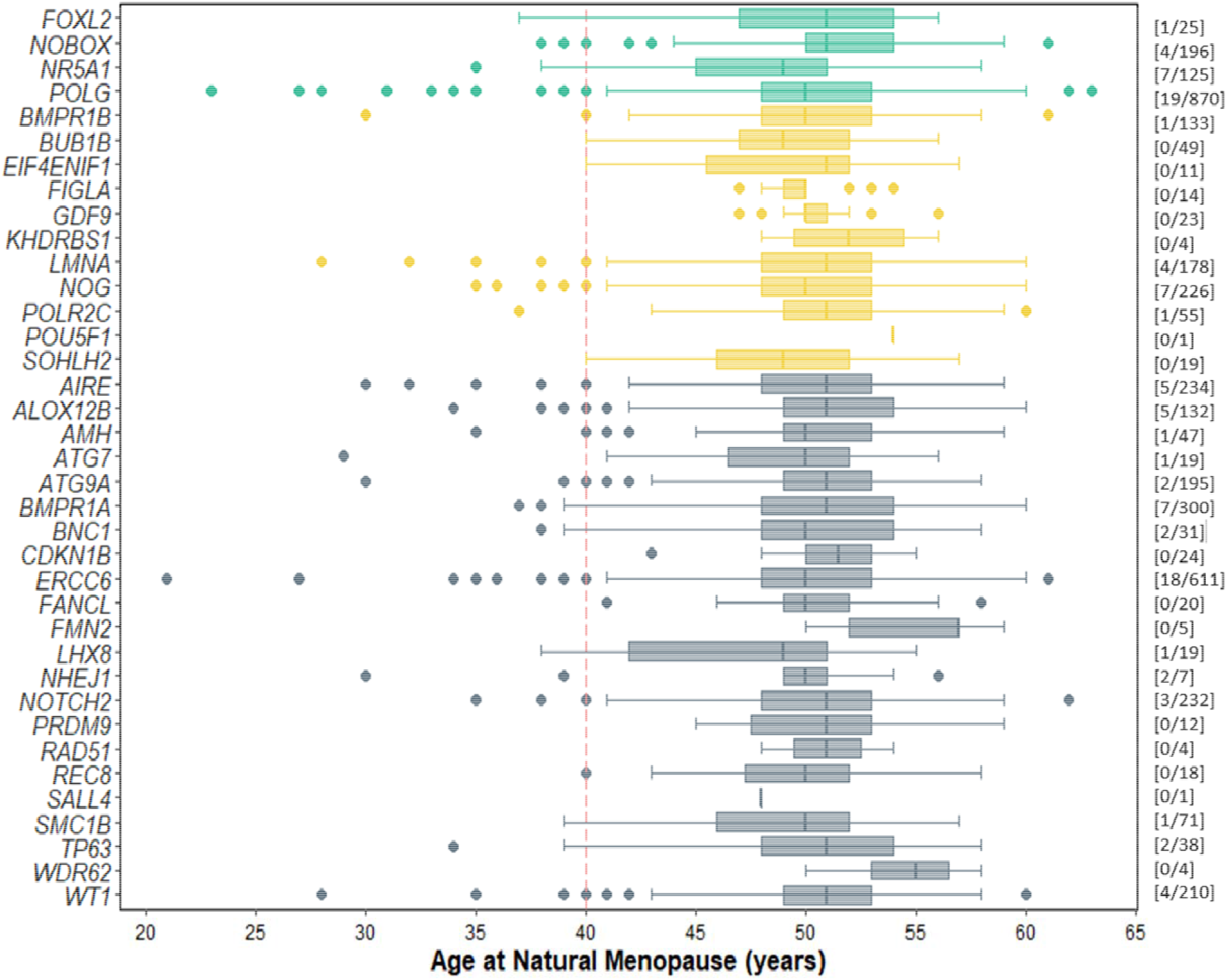
Range of age at natural menopause in carriers of missense variants with REVEL score greater than 0.7 in genes reported to have an autosomal dominant pattern of inheritance. 17 genes were identified as ‘monoallelic’ in Genomics England (GeL) Panel App and are coloured according to the strength of evidence categories: “GREEN”, and “AMBER” **(supplementary table 2)**. In addition,24 genes were reported in the literature to be a likely monogenic cause of POI in the heterozygous state but were not included on the Panel App (coloured grey). The numbers in brackets in the right corner reported as part of each panel represent **[N POI cases/N controls]** of women carrying HC PTVs in each gene. Note: In the plot, the boxes show the values of the lower quartile, median and upper quartile; the whiskers show the most extreme value within a distance of 1.5 times of the interquartile range from the lower and upper quartiles, respectively; outliers are shown as individual points.

**Supplementary Figure 3:**
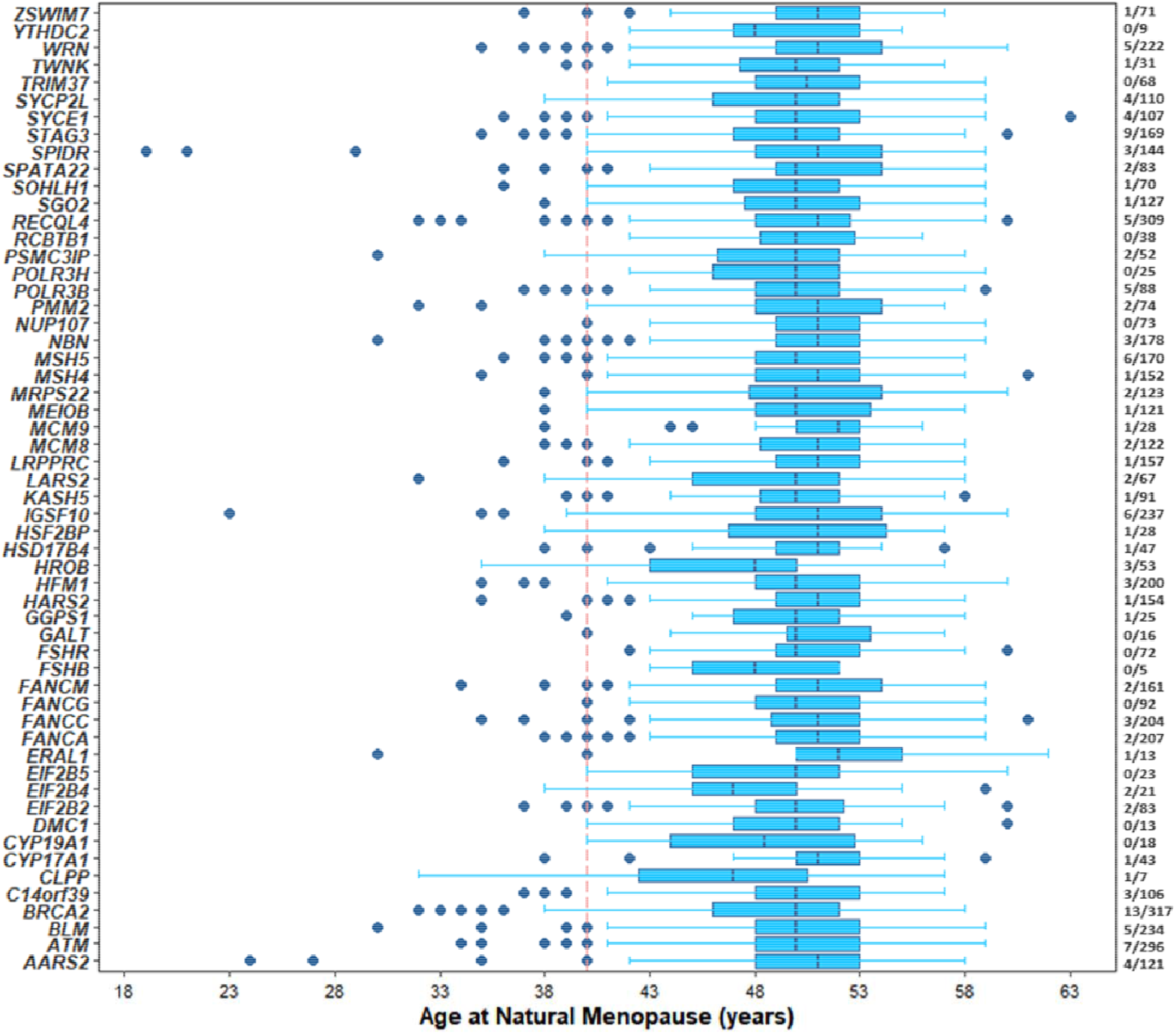
Age at natural menopause in carriers of HC-PTVs in POI genes reported to have an autosomal recessive pattern of inheritance. 65 genes were identified as ‘biallelic’ in Genomics England (GeL) Panel App **(supplementary table 2). [N POI cases/N controls]** of women carrying HC PTVs in each gene. Note: In the plot, the boxes show the values of the lower quartile, median and upper quartile; the whiskers show the most extreme value within a distance of 1.5 times of the interquartile range from the lower and upper quartiles, respectively; outliers are shown as individual points.

